# Statistical analysis plan for the STatin TReatment for COVID-19 to Optimise NeuroloGical recovERy (STRONGER) study: a randomised, open label, controlled trial in subjects with persistent neurological symptoms after COVID-19 infection

**DOI:** 10.1101/2025.07.03.25330859

**Authors:** Xiaolei Lin, Carlos Delfino, Cheryl Carcel, Paula Muñoz Venturelli, Sharon Naismith, Mark Woodward, Ruth Peters, Nirupama Wijesuriya, Meng Law, Ian H. Harding, Xia Wang, Karin Leder, Owen Hutchings, Marina Skiba, Ximena Stecher, Ella Zomer, Sophia Zoungas, Craig S. Anderson

## Abstract

We outline the statistical análysis plan (SAP) for the STatin TReatment for COVID-19 to Optimise NeuroloGical (STRONGER) study. STRONGER is an international, investigator-initiated and conducted, multicentre, prospective, randomised, open label, blinded endpoint (PROBE) study that aims to determine the safety, efficacy and cost-effectiveness of atorvastatin to improve cognitive function, mood, health-related quality of life, and physical function, in adults with a post-COVID-19 neurological syndrome. Participants are randomised (1:1) to either oral 40 mg atorvastatin for a treatment duration of 12 months (intervention group) or not (control group), on top of usual care. The treatment effect on neuroinflammation in the brain will be assessed by changes on magnetic resonance imaging (MRI) and blood biomarkers.The outcome assessments on participants will be completed in July 2025. The trial is registrated at Clinicaltrials.gov (NCT04904536).

## 1 Introduction

### 1.1 Study synopsis

Full details of the protocol for the STatin TReatment for COVID-19 to Optimise NeuroloGical (STRONGER) study have been published.^1^ In brief, STRONGER is an international, investigator-initiated and conducted, multicentre, prospective, randomised, open label, blinded endpoint (PROBE) study. The outcome assessments on participants will be completed in July 2025. Participants are randomised (1:1) to either oral 40 mg atorvastatin for a treatment duration of 12 months (intervention group) or not (control group), on top of usual care. The aim is to determine the efficacy of atorvastatin to improve cognitive impairment, mood, health-related quality of life, and physical function, in participants with a post-COVID-19 neurological syndrome through an effect on neuroinflammation in the brain indicated by changes on magnetic resonance imaging (MRI) and blood biomarkers. The trial is registrated at Clinicaltrials.gov (NCT04904536).

### 1.2 Study population

The trial recruits eligible adults (age ≥18 years) with history of COVID-19 and ongoing neurological symptoms related to impairment of memory and concentration, sleep disturbance, fatigue, or loss of smell (anosmia). Excluded people are those with dementia or significant cognitive impairment on screening, another serious health condition, history of traumatic brain injury, clear indication or contraindication for statin use, abnormal blood biochemical tests, or being female of child-bearing potential, currently breastfeeding, or planning a pregnancy. For participants who agree to undergo a brain MRI, they must have no contraindication due to metallic body parts or claustrophobia.

### 1.3 Study interventions

Eligible participants are randomised to receive treatment (oral atorvastatin 40 mg) or no treatment, on top of standard care for a period of 12 months. The randomisation is stratified by country, time since acute COVID-19 illness (<6 vs. ≥6 months), age (<60 vs. ≥60 years), current anosmia (yes vs. no), and participation in the MRI/biomarker substudy. The randomisation allocation is blinded to researchers conducting the cognitive assessments and endpoint adjudication; participants, physicians and other study team members were aware of the treatment allocation.

### 1.4 Outcomes

#### 1.4.1 Primary outcome

The primary outcome is processing speed, assessed by the oral version of the symbol digit modalities test (SDMT), whereby participants match 9 abstract symbols with numbers. Performance is measured by the number of correct symbols matched within 90 seconds.

#### 1.4.2 Secondary outcomes

Secondary outcomes include a comprehensive battery of cognitive assessments covering executive functions, memory, and other domains, alongside evaluations of health status, brain MRI and blood biomarkers. Furthermore, a cost-effectiveness analysis, relative to standard care, will be executed. Cognitive assessments are administered by trained research psychologists specialised in cognitive measures. The assessments can be conducted either in person or via videoconference. The table provides an overview of the various evaluations.

#### 1.4.3 Safety outcomes

This study documents anticipated adverse reactions to the study medication of special interest (AESI) of myalgia, nausea, elevated blood glucose, elevated creatine kinase, abdominal pain, new-onset diabetes mellitus, and rhabdomyolysis. Additionally, details are collected on any serious adverse events (SAEs) according to standard definintions, and any adverse events (AEs), in relation to severity and discontinuation of the study treatment.

## 2 Analysis principles

### 2.1 Sample size

A sample of 410 subjects was estimated to provide 80% power (α=0.05) to detect an effect size of at least a 0.3 standard deviation (SD) difference between randomised groups, on the assumption of equal group participation, 5% non-compliance and 5% dropout. The age-adjusted mean score on the SDMT is estimated at 60 (SD 13) at baseline (based on healthy control data).^2^ This effect size is based on trials of statin for the prevention of dementia and treatments for multiple sclerosis, where achieved effect sizes of 0.3-0.4 are clinically meaningful and likely to confer public health benefits.^3^ For the substudy of people undergoing MRI, a sample size of 220 (110 per group) was estimated to provide 80% power (α=0.05) to detect an effect size of relative difference of 5.0-6.5 (variance between groups/variance within groups) for the imaging endpoints (defined in the Table), on the assumption of a 20% drop out.

### 2.2 Software

Analyses will be conducted using SAS Enterprise Guide (version 8.3 or above) and R (version 4.0.0 or above).

### 2.3 Data collection

Data are compiled in a secure Web-based data management system (IBM Clinical Development) at The George Institute for Global Health. Data entry is performed at the participating sites by authorised site staff who have completed training and been given appropriate role-based access to the system. Data logic and consistency checks are programmed into the data entry forms so that data entry errors are captured in real-time and queries auto-generated. Authorised electronic signatures are used to lock completed data entry forms once all data queries have been resolved within the system. Data entry and all subsequent changes or deletions are captured in an accessible audit trail. Coding of outcomes is centrally performed either automatically via the IBM coding module or manually by the Central Coordinating Centre (CCC). All coding is reviewed and verified by a Medical Monitor. Reporting within the system is used for regular data reviews and overall trial monitoring. Data are stored and backed up on IBM cloud servers in the United States.

### 2.4 Data sets analysed

#### 2.4.1 Analysis populations

The analysis will follow the intention-to-treat (ITT) principle, whereby all patients who were randomised regardless of diagnosis and whether they received study treatment according to the protocol will be included, excluding those who withdrew their consent for any data to be used. An analysis of the per-protocol (PP) population will also be performed, comprising all patients from the ITT population with a non-missing primary outcome and who did not have a relevant protocol violation.

#### 2.4.2 Analysis strategy

Baseline characteristics will be summarized by treatment group. Continuous variables will be summarised by means (SD) if normally distributed, or medians (IQR) if normality is violated.

The primary outcome (processing speed, assessed by SDMT) will be analysed using a multiple linear regression model, where SDMT is the dependent variable, group allocation is the independent variable and study site (Melbourne, Sydney, Santiago), age and sex are included as covariates. Transformation of SDMT to approximate normality will be performed in case of severe skewness. A sensitivity analysis will use a dichotomisation of the primary outcome as high or low processing speed (SDMT <54 vs. ≥ 54 sec) in a logistic regression model with the same adjustments. Sensitivity analysis will also be conducted by including additional adjustments for year of enrolment, baseline SDMT, and level of education (4 categories of secondary or less, technical / vocational qualification, university undergraduate degree, or postgraduate degree or professional qualification).

All secondary continuous outcomes will be analyzed similary; ordinal secondary outcomes will be analyzed using cumulative logit models.

Safety outcomes will be described for each treatment arm, with continuous outcomes presented with mean (SD) or median (IQR), and categorical safety outcomes presented with frequency per category.

Subgroup analyses will be conducted on the primary outcome stratifying by the 6 baseline variables of age (<60 vs. ≥60 years), sex, time since COVID-19 diagnosis (<180 vs. ≥180 days), C-reactive protein (CRP) level (<10 mg/L vs. ≥10 mg/L), ethnicity (Caucasian vs non-Caucasian), cardiovascular risk (no vs yes, for any history of hypertension, hyperlipidaemia, current smoker, and body mass index [BMI] ≥28).

A cost-utility analysis using trial data from the EuroQoL health-related quality of life (HRQoL) EQ-5D-5L questionnaire, drug costs, and health service utilisation costs (including AEs) will be conducted, comparing use of atorvastatin and standard care. Modelling beyond the trial duration, assuming a 5-year time horizon, will also be undertaken, with analyses conducted in line with accepted standards for use of economic evaluation in decision-making.

## 3 Planned analyses

### 3.1 Subject disposition

The flow of patients through the trial will be displayed in a Consolidated Standards of Reporting Trials (CONSORT) diagram. The report will include the number of screened subjects who met the inclusion criteria and number of subjects included and reasons for exclusion of non-included subjects.

### 3.2 Patient characteristics and baseline comparisons

Baseline characteristics will be summarised by treatment group. Categorical variables will be summarised by frequencies and percentages. Percentages will be calculated according to the number of patients for whom data are available. No statistical test will be performed on baseline characteristics.

### 3.3 Analysis of the primary outcome

The primary outcome will be analyzed in a multiple linear regression model, adjusting for study site, age and sex.

#### 3.3.1 Main analysis

The main analysis will be performed in a linear regression model, with treatment allocation (atorvastatin 40 mg vs. standard care alone) included as independent variable, and study site (Melbourne, Sydney, Santiago), age and sex, included as covariates for adjustment:

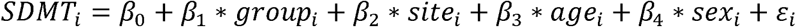

where group is coded as 1 for atorvastatin and 0 for standard of care group, respectively.

The effect of the intervention will be presented as the difference of SDMT between atorvastatin group and standard care group (i.e., estimate of *β*_1_ in the regression model). The associated 95% confidence intervals (CI) can be calculated by (*β̂*_1_ - 1.96 * *sê*, *β̂*_1_ + 1.96* *sê*), where *sê* is the estimated standard error of *β̂*_1_. A difference of SDMT >0.4 corresponds to a clinically better outcome for atorvastatin group compared to standard care alone group.

If the distribution of SDMT is highly skewed, we will perform transformation (log or square root transformation) to SDMT before conducting a linear regression analysis.

If the estimated difference of SDMT between atorvastatin and standard care group exceeds 0 and 95% CI does not include 0, then we conclude that atorvastatin plus standard care improve processing speed compared to standard care alone with statistically significance. If the estimated difference of SDMT between atorvastatin and standard care group exceeds 0.4 and 95% CI does not include 0.4, then we conclude that atorvastatin plus standard care improve processing speed compared to standard care alone with clinical significance.

#### 3.3.2 Subgroup analyses

Six pre-specified subgroup analyses will be carried out, irrespective of whether there is a significant treatment effect on the primary outcome. These subgroup analyses will only be performed in the ITT population.

Subgroups based on patient characteristics assessed before randomisation are defined as follows:

- age (< 60 vs. ≥60 years);
- Sex (female vs male)
- time since COVID-19 diagnosis (<180 days vs. ≥180 days);
- baseline CRP levels (<10 mg/ L vs. ≥10 mg/ L);
- ethnicity (Caucasian vs. non-Caucasian);
- prior cardiovascular risk (no vs yes, for any history of hypertension, hyperlipidaemia, current smoker, and high BMI);

The analysis for each subgroup will be performed by first stratifying the ITT population according to each subgroup variable, and then calculating the difference of SDMT (and 95% CI) within each stratum. The results will be displayed on a forest plot, including the point estimate and 95% CI associated with the SDMT difference in each stratum of the subgroup.

#### 3.3.3 Treatment of missing data

The proportion of data missing for the primary outcome (due to participants lost to follow-up) will be described. In the case of a non-negligible amount of missing data (>5%), we will use multiple imputations by chained equation to assess the impact of missing data to the results.^4,5^ If proportion of missingness is less than 5%, we will use complete data for analyses.

#### 3.3.4 Sensitivity analyses

The analyses of the primary outcome will be conducted by dichotomizing SDMT by 54 (the normative median score in the general Australian and Chile population) and by using logistic regression, where study site (Melbourne, Sydney, Santiago), age and sex are included as covariates to be adjusted. Code partipants with SDMT <54 as having y=0, while those with SDMT ≥54 as y=1, the logistic regression model can be represented by

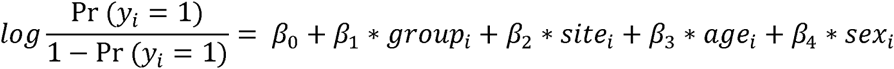

where group=1 indicates a participant is allocated to the atorvastatin group, and group=0 indicates a partipant is allocated to the standard care group. The parameter *β*_1_ will capture the adjusted treatment effect associated with atorvastatin. The point estimate of the treatment effect (odds ratio [OR]) is estimated by e^*β̂*_1_^ (exponential of the maximum likelihood estimate of *β*_1_), and 95% CI can be obtained by (e^(*β̂*_1_-1.96\**sê*)^, e^(*β̂*_1_+1.96\**sê*)^), where *se* is the estimated standard error of *β*_1_. Results will be presented by the estimated OR (high SDMT vs. low SDMT) of atorvastatin relative to standard care group, with estimated 95% CI. If the estimated OR of SDMT of atorvastatin relative to standard care group exceeds 1 and 95% CI does not include 1, then we conclude that atorvastatin plus standard care improve processing speed compared to standard care alone with statistical significance.

Additional sensitivity analyses will be conducted by (i) modelling SDMT with additional adjustments for year of enrolment, baseline SDMT, and level of education (4 categories), and; (ii) using other approaches to handle missingness, such as mean imputation.

The main analyses and the subgroup analyses will be repeated in the PP population.

### 3.4 Analysis of secondary outcomes

All secondary outcome analyses will be performed in the ITT (primary) and PP (sensitivity) population. Secondary outcomes will be grouped into 4 families, as outlined below.

**Cognitive family:** *Learning*, assessed by RAVLT15 raw score; *Memory*, assessed by RAVLT7 raw score; *Executive*, assessed by CWIT raw score (note: age scaled scores are converted to z-scores); *Executive functioning composite*, assessed by the average z-score of semantic fluency (SF), CWIT3, Trails B if z-scores available; *Global composite*, assessed by the average z-score of TMTA, CWIT1, RAVLT15, RAVLT7, Semantic fluency, TMTB and CWIT3.

**Health assessment family:** *Overall health*, assessed on the COVID-19 Yorkshire Rehabilitation Scale; *Depression*, assessed by Patient Health Questionnaire (PHQ-9); *Anxiety*, assessed by General Anxiety Disorder (GAD-7); *Sleep*, assessed on the Pittsburgh Sleep Quality Index (PSQI); *HRQoL*, assessed by EQ-5D-5L on each of the 5 domains, visual analogue scale (VAL), and overall health utility; *Physical activity*, on the International Physical Activity Questionnaire- Long Form (IPAQ-LF)

**Brain imaging family:** *Mean white matter free water fraction (primary)*, total volume of perivascular spaces, total grey matter volume and average cortical thickness, mean white matter microstructural integrity (fractional anisotropy), total white matter hyperintensity volume, mean cerebral perfusion, and mean basal ganglia iron load.

**Blood biomarker family:** NFL protein, TNF-a, GFAP, Ptau-181, Aβ42/40, DNA extraction for apolipoprotein E genotype, IL-6, IL-1β, NAD+, hsCRP.

Sensitivity analyses of the brain imaging outcomes will be conducted by using regional brain volumes.

#### 3.4.1 Continuous secondary outcomes (RAVLT-D, OTMT-A, OTMT-B, D-KEF CWIT, SF, IPAQ-LF)

These will be analysed using linear regression, with continuous secondary outcome included as the dependent variable, group included as the independent variable, and study site (Melbourne, Sydney, Santiago), age and sex, included as the covariates to be adjusted.

#### 3.4.2 Categorical secondary outcomes (PHQ-9, GAD-7, PSQI, IPAQ-LF)

Binary secondary outcomes, including PHQ-9 (depression vs no depression) and PSQI (poor sleep vs good sleep), are analyzed using logistic regression, where binary secondary outcome is included as the dependent variable, group is included as the independent variable, and study site (Melbourne, Sydney, Santiago), age and sex, are included as the covariates to be adjusted. Multi-category secondary outcomes, including GAD-7 (mild, moderate, and severe anxiety) and IPAQ-LF (high, medium, low physical activity) are analysed with cumulative logit regression, and proportional odds assumption will be tested. Multi-category secondary outcome is included as the dependent variable, group is included as the independent variable, and study site (Melbourne, Sydney, Santiago), age and sex, included as the covariates to be adjusted.

If proportional odds assumption holds, the effect of the treatment can be presented as the cumulative OR (severe versus mild + moderate anxiety, severe + moderate versus mild anxiety; high versus medium + low physical activity, high + medium versus low physical activity) between treatment and control group, with 95% CI. An OR greater than 1 corresponds to an increase of anxiety level or physical activity level in the treatment group compared to the control group.

If proportional odds assumption does not hold, the effect of the treatment will be presented as OR separately for severe versus moderate + mild anxiety, and severe + moderate versus mild anxiety (or separately for high versus medium + low physical activity, high + medium versus low physical activity). An OR greater than 1 corresponds to an increase of anxiety level or physical activity level in the treatment group compared to the control group.

#### 3.4.3 Substudy outcomes (MRI markers, neurodegenerative markers, peripheral markers)

Analysis of the substudy outcomes will be performed in participants in the substudy undergoing MRI. MRI markers, including mean white matter free water (FW) fraction from diffusion MRI (dMRI), white matter hyperintensity (WMH) volume, enlarged peri-vascular space (PVS) volume, total grey matter volume, mean white matter microstructural integrity (fractional anisotropy), basal ganglia iron load and mean cerebral perfusion will be analysed with linear regression, with MRI marker included as the dependent variable, group included as the independent variable, and study site (Melbourne, Sydney, Santiago), age, sex, and total intra-cranial volume (for volumetric outcomes) included as the covariates to be adjusted. Continuous neurodegenerative markers (Ptau-181, neurofilament light chain (NfL) and Aβ42/40), and continuous peripheral markers (IL-6, IL-1β,NAD+, TNF-α, hsCRP), will be analysed with linear regression, with marker included as the dependent variable, group included as the independent variable, and study site (Melbourne, Sydney, Santiago), age and sex, included as the covariates to be adjusted.

In addition, neurodegenerative and peripheral markers that have clear clinical cutoffs will be categorized into categorical markers (such as normal versus abnormal, or low, normal, high levels). Then, binary or ordinal logistic regression will be conducted to analyze these categorical markers.

### 3.5 Analysis of exploratory outcomes

Exploratory outcomes, including employment status, household income, readmission to hospital, will be analyzed with linear regression, with exploratory outcome included as the dependent variable, group included as the independent variable, and study site (Melbourne, Sydney, Santiago), age and sex, included as the covariates to be adjusted. Analyses are considered exploratory and multiplicity will not be adjusted.

### 3.6 Analysis of SAEs and deaths

SAEs will be summarised as the number of events as well as the number and proportion of patients experiencing at least one SAE event. This will be done overall and by category of event according to Medical Dictionary for Regulatory Activities (MeDRA) system organ classes and preferred terms. Primary and underlying causes of deaths will be summarised by treatment arm with no formal test. A listing of all SAEs will be compiled (in an appendix).

### 3.7 Economic evaluation

#### 3.7.1 Overview

The primary objective of the health economic evaluation is to calculate the cost-effectiveness of atorvastatin therapy versus standard care alone over the trial duration using a within-trial economic evaluation study design. The second objective is to build an economic model to extrapolate the longer-term costs and benefits of atorvastatin therapy beyond the trial period over 5-years. A health care system perspective will be adopted as the primary perspective. A partial societal perspective, which includes impacts on return to work, will be undertaken as a secondary analysis. The main outcome of interest will be the incremental cost-effectiveness ratio (ICER) in terms of cost per quality-adjusted life year (QALY) gained. Economic evaluations typically use the outcome metric of QALYs gained because cost-effectiveness ratios using QALYs have inherent value-for-money connotations. Costs and outcomes will be presented as undiscounted and discounted in line national with recommendations. Cost-effectiveness will be determined by benchmarking the ICER/s against relevant willingness-to-pay thresholds, which represent the maximum amount a society or decision-maker is willing to pay for a unit of health gain.

The analyses will follow the Consolidated Health Economic Evaluation Reporting Standards (CHEERS) 2022 guidelines^6^ and ISPOR best practice modelling guidelines^7^ for reporting with a format appropriate to stakeholders and policy makers.

#### 3.7.2 Costs

Health care costs include the costs of atorvastatin therapy and healthcare use (e.g. admissions to hospital, other medications) utilised by study participants during the study period. All healthcare use will be derived from trial data and valued in monetary terms using appropriate unit costs. The average per patient healthcare use will be tabulated by randomised treatment group, with precision estimates (i.e., SD and 95% CI). The total cost for each participant will be calculated as the sum of atorvastatin and other concurrent healthcare costs over the study period. The partial societal perspective also incorporates effects on productivity in terms of impacts on return to work. The human capital approach will be used to value lost paid productivity using national average wages for job categories reported by study participants.

#### 3.7.3 Outcomes

The primary outcome measure will be QALYs. Utility values will be derived from participant responses at each time point on the EQ-5D-5L quality of life instrument. The utility values recorded at baseline and follow-up will be used to calculate total QALYs for each participant using the area under the curve method,^8^ adjusting for any imbalances in baseline EQ-5D-5L values.

#### 3.7.4 Statistical analysis of economic data

Mean differences in costs, QALYs and net benefits, between the randomised groups will be estimated with associated 95% CI. All statistical tests will be two-sided, and the statistical significance level will be set at <5%.

The statistical analyses for the economic evaluation will follow the principles detailed previously for the primary analysis and will employ an ITT approach, where all individuals randomised will be included in the analysis by their allocated trial arm status regardless of whether they received all, part or none of the intended treatments.

For the base case, generalised linear models (GLM) using a gamma family and log link will be used to estimate the difference in the total health sector costs between the atorvastatin and standard care groups over the trial period due to the typically right skewed distribution of cost data. The mean difference in QALYs will be estimated using GLM with the gaussian family and identity link with adjustment for the baseline covariates sex, age and utility. Separate GLMs will be used to estimate the difference in total societal costs and QALYs between treatment groups. Negative binomial regression will be used to examine the between group differences for the components of total cost related to medications, hospital visits, and lost productivity.

In addition to reporting descriptive statistics and differences between treatment groups for costs and outcomes, ICERs will be calculated as the mean difference in total cost divided by the mean difference in outcome (i.e. QALYs) between the trial arms. The 95% CI around ICERs will be calculated using a nonparametric bootstrap procedure, with 1,000 iterations to reflect sampling uncertainty.

#### 3.7.5 Sensitivity analyses

Sensitivity analyses will be conducted using complete-cases only using GLM, with and without adjustment for covariates. Complete cases will be records with HRQoL data and resource use data observed over the trial duration. The results for complete cost and health outcome data (i.e., those with no missing data) as well as a strict per-protocol analysis of the data will be provided to identify the impact of missing data on the analysis and any sensitivity to protocol violations.

#### 3.7.6 Modelled economic evaluation

The costs and outcomes data from the within trial evaluation will be used to estimate the cost-effectiveness of statin therapy over a 5-year time horizon using economic modelling techniques. More details regarding the modelled economic evaluation will be provided after the within trial economic evaluation has been completed. The modelling will only be undertaken if atorvastatin is found to be effective.

## Data Availability

Individual, de identified participant data used in these analyses may be able to be shared on request from any qualified investigator following approval of a protocol and signed data access agreement via both the trial steering committee and the Research Office of The George Institute for Global Health, Australia.

## Author contributions

XL, CC, PMV, SN, MW, RP, NW, ML, IHH, XW, KL, OH, MS, XS, EZ, SZ and CSA made substantial contributions to the conception and the design of the work. XL, EZ and CSA, wrote the first and following drafts of this manuscript. All other authors contributed to the critical revision of manuscript for intellectual content. All authors approved submission of the manuscript for publication.

---

**Table.**
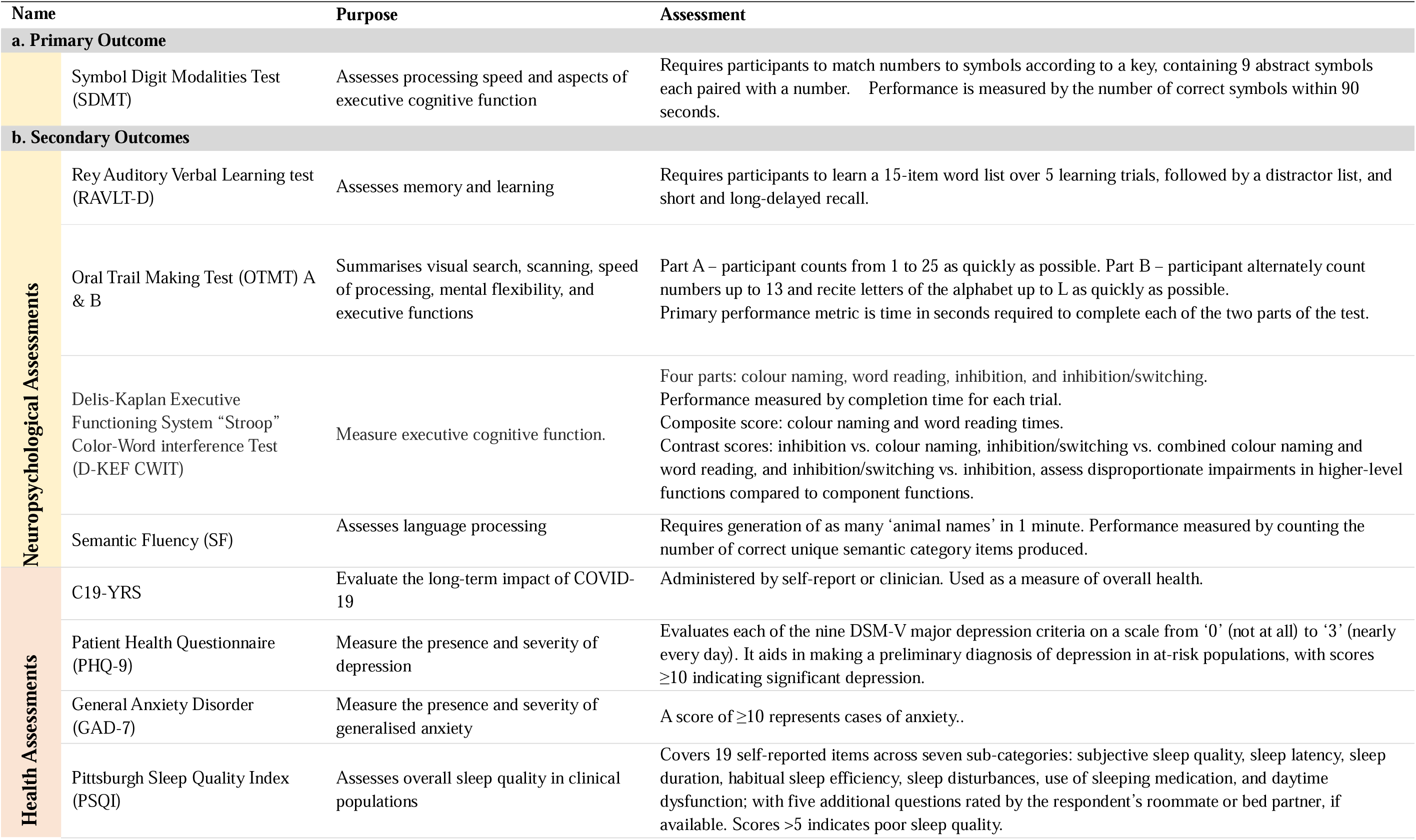
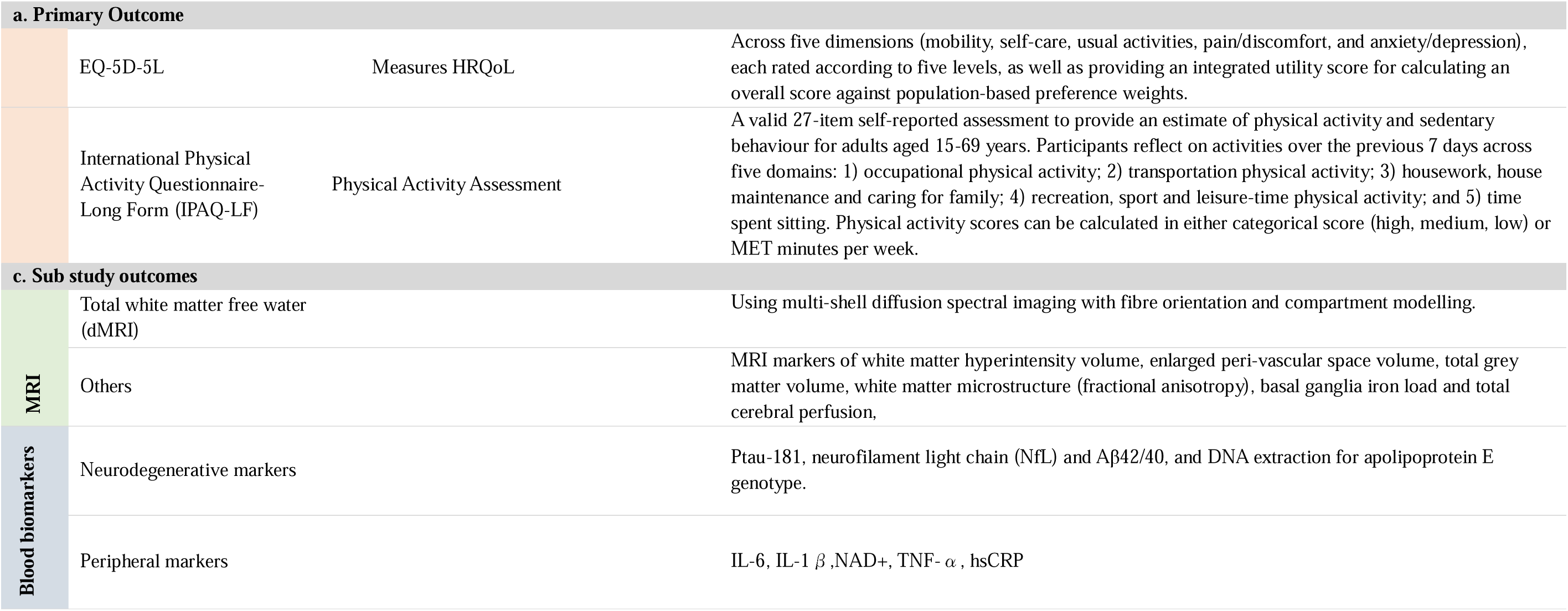
Study Outcomes.

## 3 Appendix: Proposed tables and figures

**Table 1:**
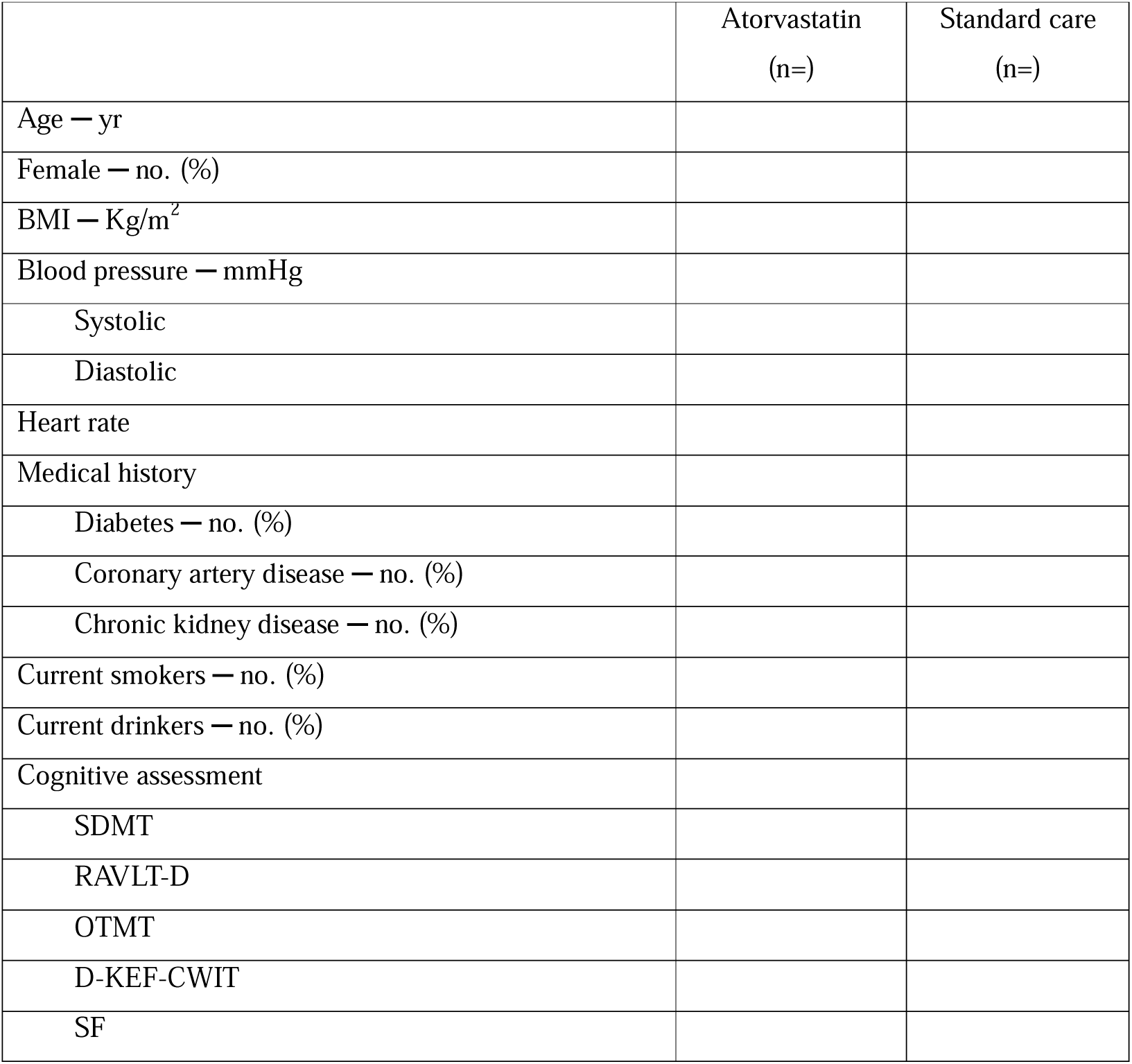
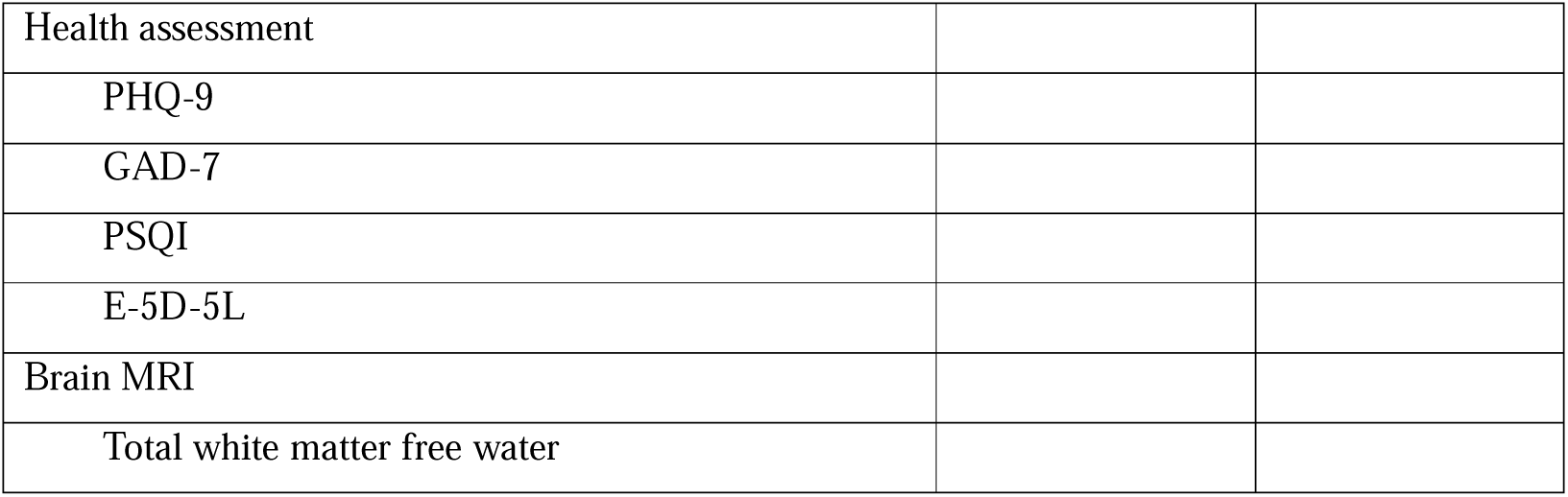
Baseline Characteristics.

**Table 2:**
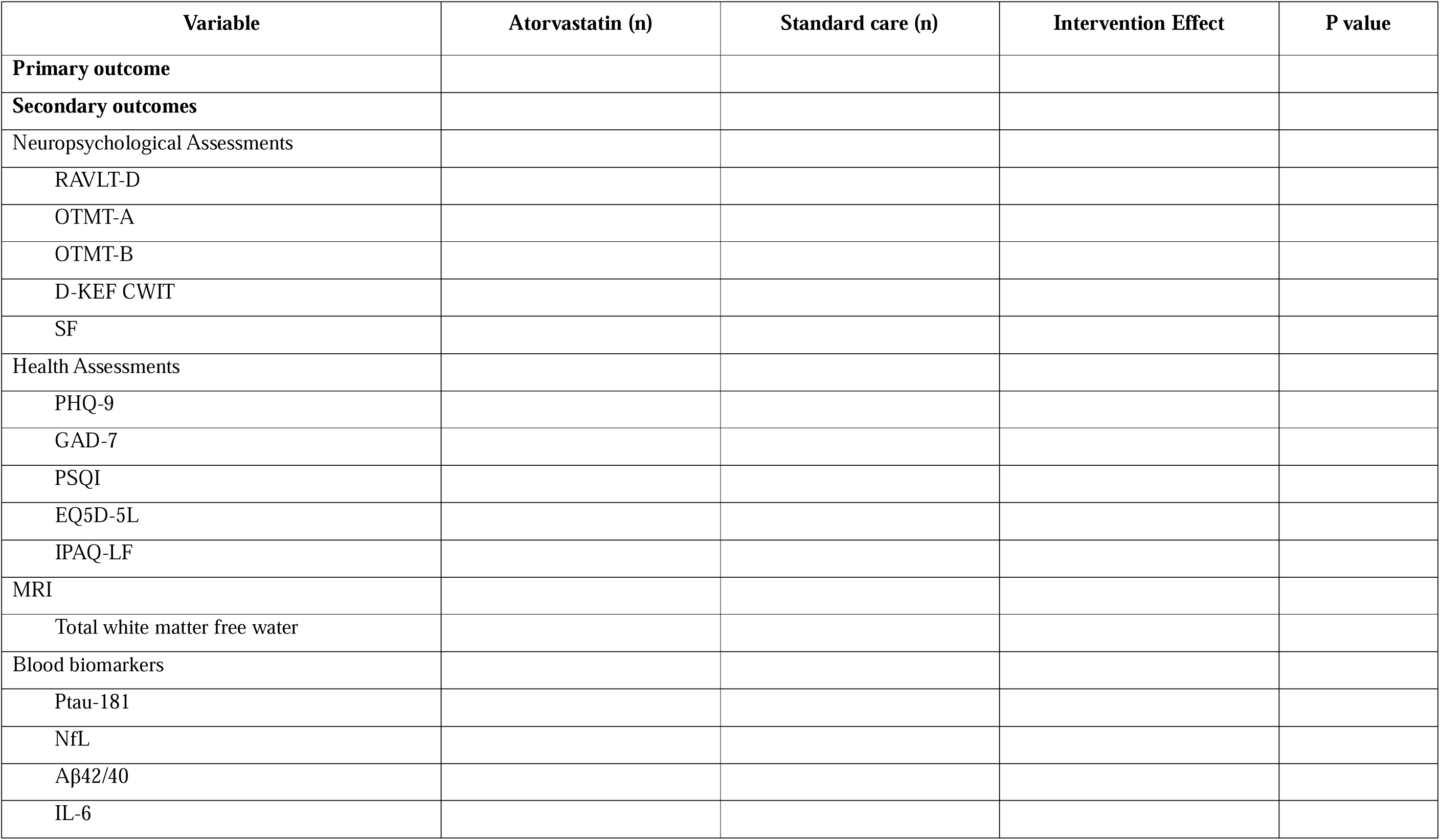
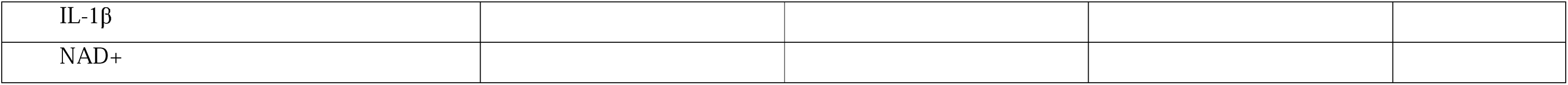
Intervention Effect on Primary and Secondary Outcomes.

**Table 3:**
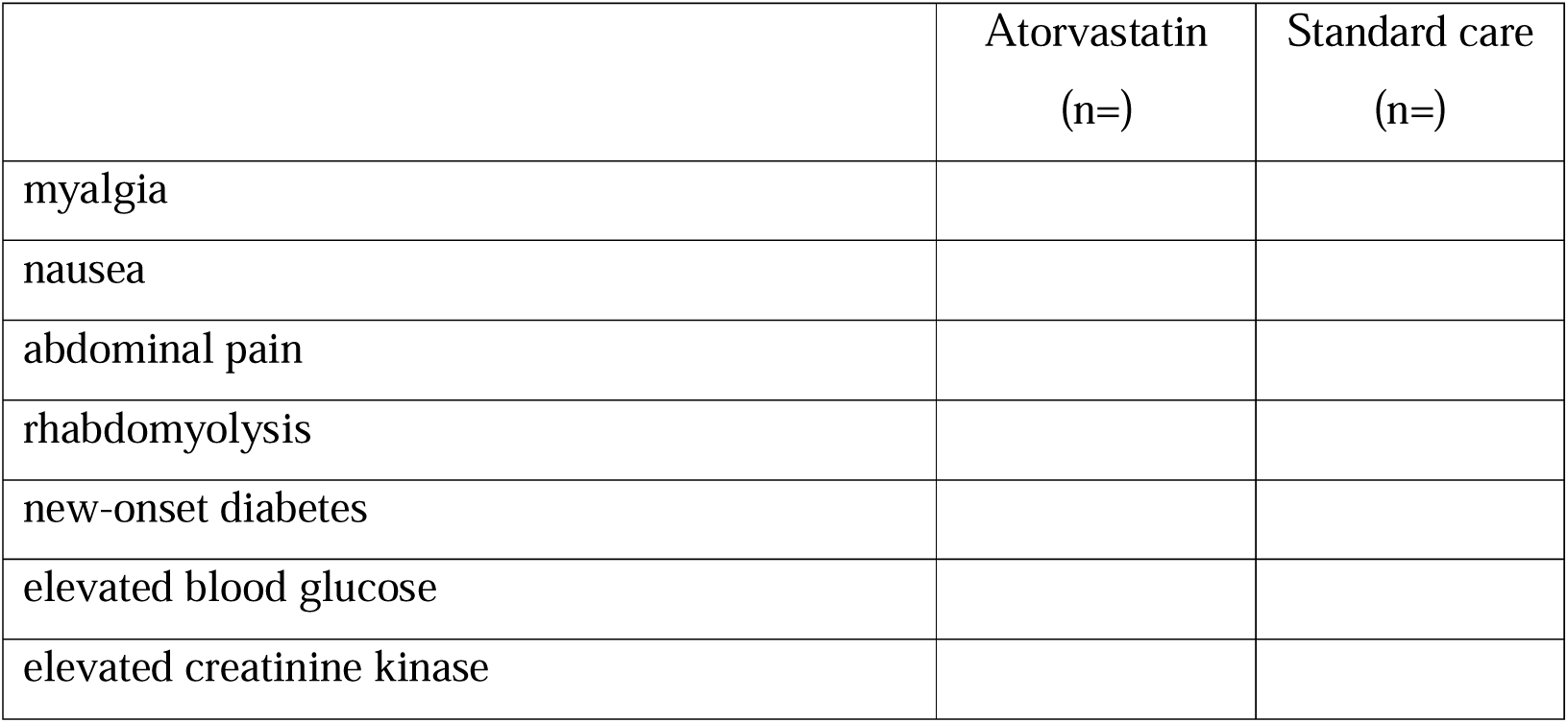
Description of AESIs.

**Figure 1:** Flow diagram of participants.

**Figure 2:** Distribution of SDMT in the atorvastatin and standard care group.

**Figure 3:**
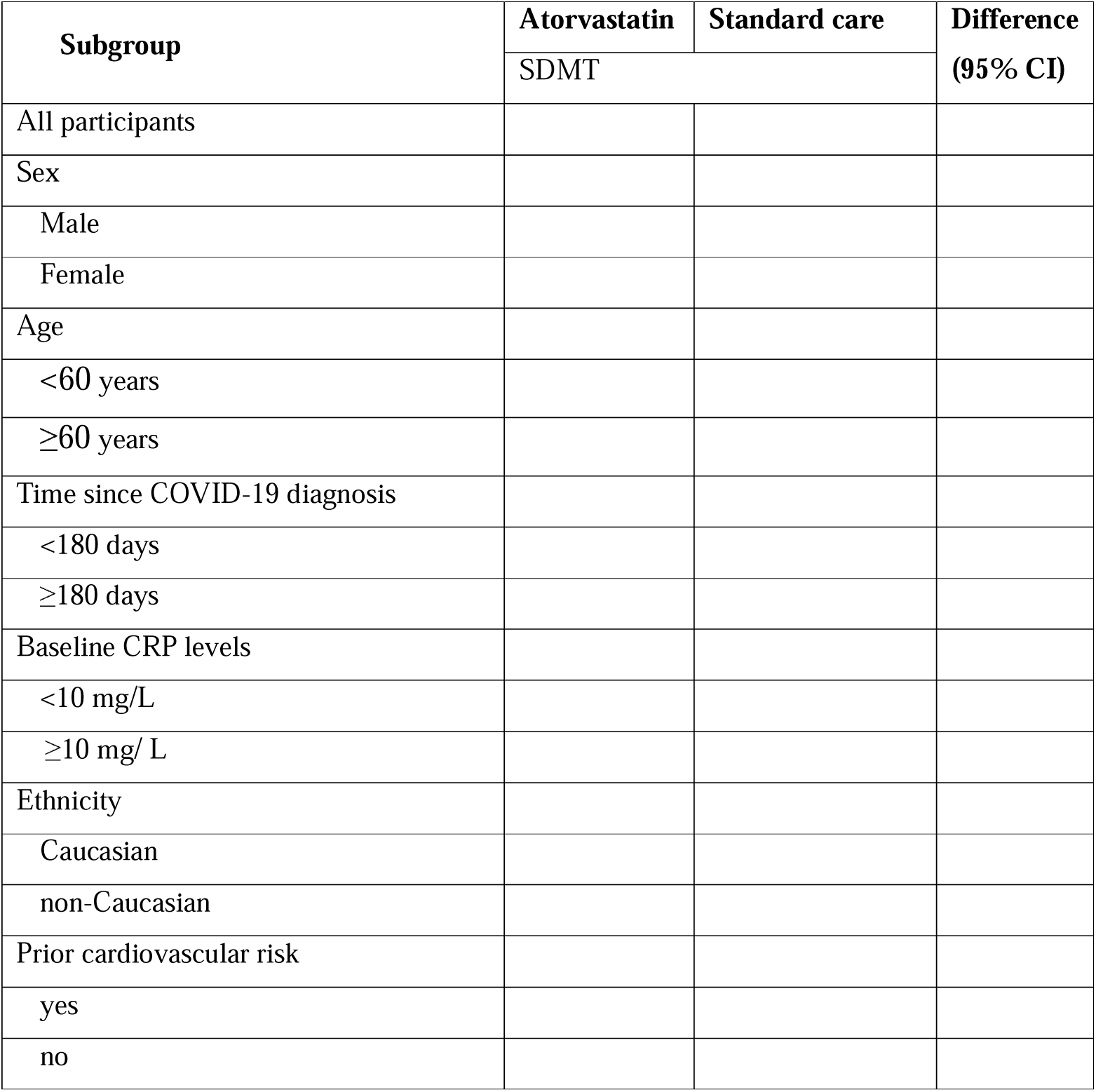
Subgroup analyses for SDMT.

